# A Phylogeny-Informed Mathematical Modeling of H5N1 Transmission Dynamics and Effectiveness of Control Measures

**DOI:** 10.1101/2025.07.29.25331913

**Authors:** Oluwatosin Babasola, Mohamed Bakheet, Christina Naesborg-Nielsen, Sachin Subedi, MHM Mubassir, Justin Bahl

**Affiliations:** College of Veterinary Medicine, Department of Infectious Diseases, University of Georgia, Athens, GA; Center for Ecology of Infectious Diseases, University of Georgia, Athens, GA; Institute of Bioinformatics, University of Georgia, Athens, GA; Department of Epidemiology and Biostatistics, University of Georgia, Athens, GA

**Keywords:** Bird flu, Control measure, Optimal control, Phylogeny, Stability analysis, Vaccination, Zoonotic transmission

## Abstract

The highly pathogenic avian influenza (HPAI) subtype H5N1 is a severe viral disease which continues to pose a significant threat to public health and a rigorous understanding of its transmission dynamics across its major pathways is essential for developing effective control strategies. Phylogenetic analysis suggests that H5N1 spillover occurs primarily between wild and domestic birds. However, increasing contact between these species and humans increases the risk of zoonotic transmission. In this work, we develop a mathematical model to examine the transmission dynamics of H5N1 and evaluate the effectiveness of proposed control measures. The model employs a compartmental framework that includes human, domestic, and wild bird populations. We then use this model to estimate the basic reproduction number for each population group and perform a sensitivity analysis to assess the contribution of the parameters to the spread of the disease. Numerical simulations are also conducted to evaluate the impact of inter-species interactions on H5N1 infection in humans and to determine the effectiveness of different control measures. The results suggest that a vaccination strategy with high vaccine efficacy, combined with a vaccination rate above 50%, significantly reduces transmission. In addition, decreasing cross-species interactions leads to a substantial reduction in disease transmission within the human population. Moreover, an optimal control analysis indicates that a combined approach involving environmental sanitation, vaccination, and targeted culling of confirmed infected poultry is an effective strategy to control the outbreak, reducing the likelihood of spillover to humans.

## 1. Introduction

Highly pathogenic avian influenza subtype H5N1 is a known pathogen that causes infection in both domestic and wild birds [1]. However, sporadic spillover events have occurred, with the current strain infecting not only avian hosts but also a range of mammalian species [2–6]. Recently, multiple cases have been reported worldwide in cattle, domestic cats, alpacas, wild raccoons, and humans [7]. For instance, in the United States, the H5N1 outbreaks have been recorded in poultry and various mammalian species, with a few human infections linked to direct exposure from infected dairy cattle and poultry [8]. According to the Centers for Disease Control and Prevention (CDC) [1], nearly all human H5N1 cases prior to 2024 were associated with direct contact with infected poultry and till date, no human-to-human transmission has been recorded. However, the possibility of future adaptation cannot be ignored as it has been reported in multiple sources that avian influenza virus has an affinity for human cells [9–13]. This raises serious concerns about the virus’s potential capability of sustaining human-to-human transmission. As a result, it is essential to fully investigate and understand the possible transmission pathways and identify the risk of human infection with the goal of developing mitigation approaches. Numerous studies have explored the transmission dynamics of H5N1 and strategies to mitigate its spread [2, 9, 14, 15]. Despite these efforts, the virus remains a public health threat and poses a significant risk to the global health and economy [16–18]. To provide improved understanding of the virus transmission risk in humans and potential control measures, we combined phylogenetic analysis and mathematical modeling approaches. The mathematical models have proven to be a valuable tool for understanding infectious disease dynamics and it will be essential in examining the risk of transmission of H5N1 from domestic and wild bird hosts to humans. In fact, several studies have adopted mathematical approaches to examine the spread of the virus within poultry populations. For instance, Malek et al. [19] developed a deterministic six-compartment model incorporating vaccination and treatment in poultry farms with the assumption that poultry remains the natural host of the virus. Through the numerical simulations, their result suggests that outbreak size is influenced by vaccine availability and treatment rates. Similarly, Kharis et al. [20] analyzed the impact of bird vaccination in a constant population. They identified the significant parameters responsible for rapid transmission and then performed a theoretical analysis of the influenza epidemic. Even though much work has been done in understanding transmission dynamics of H5N1, many of the existing models only focus on the within-species transmission, while cross-species interactions are not well accounted for. Given recent evidence of H5N1 transmission from wild birds to poultry and from infected cattle to humans, a more comprehensive approach is needed. This work develop a mechanistic model for the cross-species transmission dynamics of H5N1 among wild birds, domestic birds, and humans. To this end, we formulate a transmission dynamics model of avian influenza in different species which will enable us to model the interactions between the populations. Without loss of generality, we limit the ambiguity of this model by categorizing the model into three groups: human (*h*), domestic birds (*d*) with a focus on poultry, and wild birds (*w*). The remainder of this article is structured as follows: Section 2.1 outlines the model formulation and parameter definitions. Section 2.2 presents the model analysis, including the fundamental properties and the estimation of the basic reproduction number. Section 2.6 focuses on model fitting and parameter estimation, while Section 3.1 provides numerical simulations to assess the impact of cross-species transmission on human infections. In Section 3.2, we extend the model to incorporate optimal control strategies and evaluate intervention strategies. Finally, Section 4 concludes the article with a summary of key findings and recommendations for future work.

## 2. Methods

### 2.1. Model Formulation

We designed our model to comprise of three population hosts that were further sub-grouped into different compartments based on their epidemiological status. The human and domestic population were modeled using the SVEIR compartmental model, while the wild population follows the SI compartmental model. We denote the susceptible population as *S*_*i, i*=*h,d,w*_, which comprises of individuals who are not yet exposed to the virus but are at risk of becoming exposed and infected after an effective contact with the virus. The vaccinated population *V*_*h,d*_ represents the proportion of susceptible individuals who have received a vaccine to reduce their likelihood of infection after exposure. Individuals in this group are expected to have a level of immunity to the virus depending on the vaccine efficacy. According to [21, 22], the U.S. Department of Health and Human Services (HHS) has awarded a pharmaceutical company (Moderna) the sum of $590 million to develop a human vaccine against avian influenza, which is in addition to the $176 million previously allocated for early-stage research. This gives a clear indication that the vaccination strategy could be adopted in the future. The exposed group *E*_*j, j*=*h,d*_ includes the population that has been in contact with the virus but is not yet infectious. Then we have the infected population denoted as *I*_*i, i*=*h,d,w*_, which consists of individuals capable of transmitting the virus through direct or indirect contact with the susceptible group. Next, we define the recovered population *R*_*j, j*=*h,d*_. This population includes individuals who have survived infection through an intervention strategy or natural immune response. It is important to note that only the human and domestic population groups have the vaccinated compartment, while it is omitted within the wild birds population. This omission is mainly due to the impracticality of vaccinating wild birds on a large scale. As noted in [23], the implementation of a wild bird vaccination strategy presents a significant challenge, as it is not feasible to capture and administer vaccines to free-ranging populations. Unlike human and domestic bird population, which can be managed within relatively controlled environments, wild birds often migrate across vast distances, making widespread vaccination unattainable. Thus, using the information above, we have 12 compartmental variables which are given in Table 1.

**Table 1:** Description of the *H*5*N* 1 model variables.

| Variables | Description |
| --- | --- |
| $S_h$ | Susceptible human |
| $V_h$ | Vaccinated human |
| $E_h$ | Exposed human |
| $I_h$ | Infected human |
| $R_h$ | Recovered human |
| $S_d$ | Susceptible domestic bird |
| $V_d$ | Vaccinated domestic bird |
| $E_d$ | Exposed domestic bird |
| $I_d$ | Infected domestic bird |
| $R_d$ | Recovered domestic bird |
| $S_w$ | Susceptible wild bird |
| $I_w$ | Infected wild bird |

For the model parameters, the susceptible population has a recruitment rate {*θ* = *θ*_*h*_, *θ*_*d*_, *θ*_*w*_}. The human susceptible population is expected to increase by the vaccine waning rate *ψ*_*h*_ and decrease as the population becomes vaccinated at the rate *ω*_*h*_. Moreover, after effective contact with an infected host, the susceptible population is expected to be reduced by the force of infection rate *ϕ*_1_ which is defined as

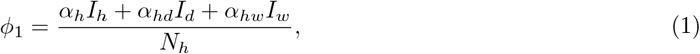

The parameters *α*_*h*_, *α*_*hd*_ and *α*_*hw*_, represent the effective transmission probability per contact of a human with human, domestic and wild respectively. Lastly, all human populations are reduced by natural death at a rate *µ*_*h*_. For the vaccinated population, this is expected to increase as the population becomes vaccinated, while it will decrease by natural death rate and the vaccine-waning rate. Furthermore, since no vaccine has 100% efficacy [24], this suggests that an individual vaccinated against H5N1 can still be infected and transmit the disease. Thus, the vaccinated population would be reduced after effective contact with H5N1 infection with a rate (1 − *γ*_*h*_)*ϕ*_1_, where *γ*_*h*_ represents the vaccine efficacy. Also, the vaccinated population is reduced by natural death of humans at a rate *µ*_*h*_. Similarly, for the exposed human population, the group would increase as the susceptible individuals have contact with the infected group and are further depopulated by the progression rate from the exposed state to the infectious state (denoted by *τ*_*h*_) due to the incubation period. According to the American Medical Association (AMA) [25], the available data suggest that the estimated incubation period for human infection with the H5N1 virus is generally 3 − 5 days, but it has been reported to be up to 7 −10 days after which an individual is expected to be symptomatic. The infectious population increases with the progression rate of exposed individuals and is reduced due to recovery from infection at a rate *σ*_*h*_, disease-induced mortality rate *δ*_*h*_ and the natural death rate *µ*_*h*_. Finally, the recovered human population (*R*_*h*_) contains individuals who have been cleared of the disease after a period of time. This population is expected to increase with the number of infectious humans that recover from infection at a rate *σ*_*h*_ but can also be reduced by the natural death rate.

Following a similar approach, the domestic group is classified as susceptible, vaccinated, exposed, infected, and recovered, while the wild population is classified as susceptible and infectious only. We note that this group follows a similar interpretation as presented in the human population, with slight differences in notation. The force of infection, for the domestic and wild groups, are denoted by *ϕ*_2_ and *ϕ*_3_ respectively, which are expressed below.

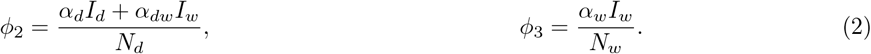

Here, the parameter *α*_*d*_ represents the effective transmission probability per contact with the infected domestic group, *α*_*dw*_ represents the effective transmission probability per contact between domestic and infected wild birds, while *α*_*w*_ is the effective transmission probability per contact between wild birds. This compartmental division is also supported by the phylogenetic analysis as shown in Figure 2.1, which involves the analysis of viral genomic sequence data for H5Nx that suggested multiple transmission pathways, observed primarily in wild, domestic birds, and humans.

**Figure 1:**
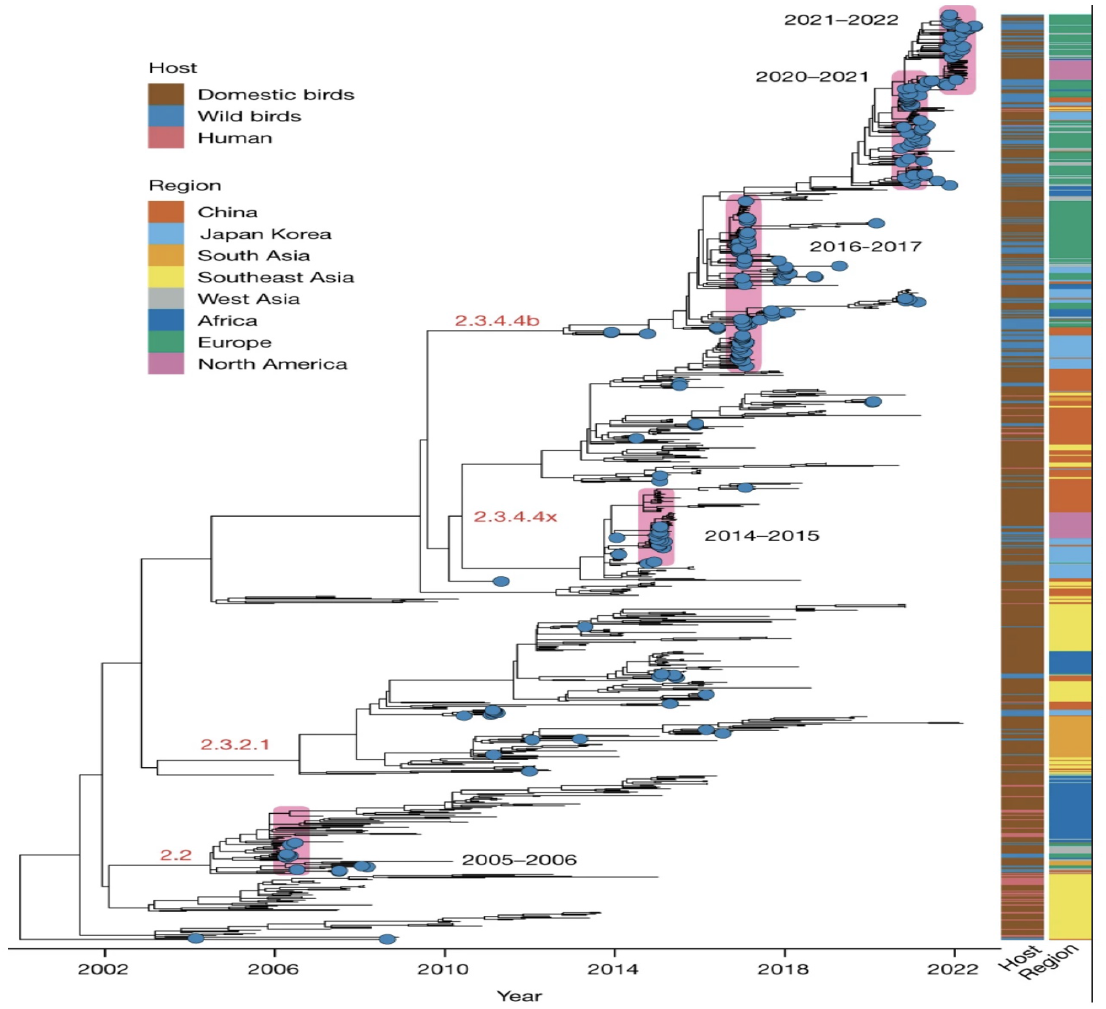
Time-scaled maximum-likelihood tree of HPAI H5 [26].

**Figure 2:**
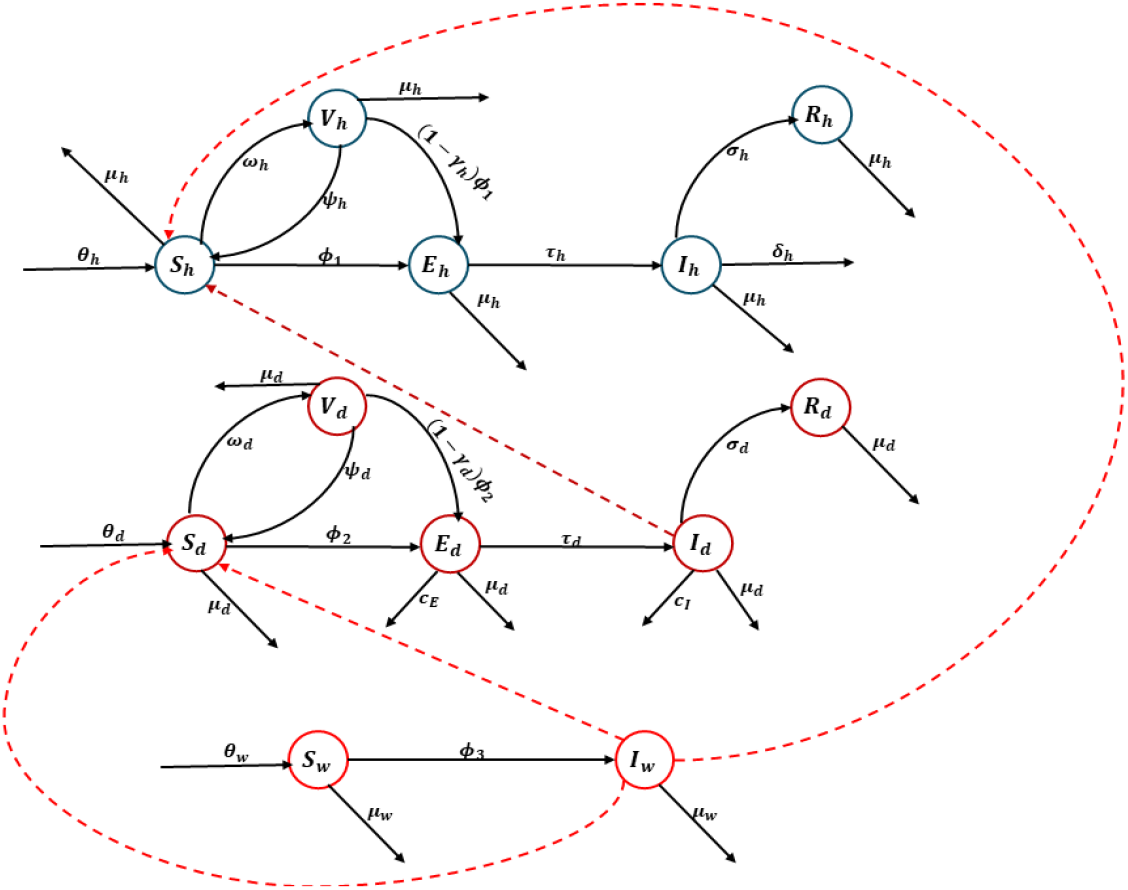
The schematic diagram representing the flowchart of the *H*5*N* 1 transmission model (4).

**Figure 3:**
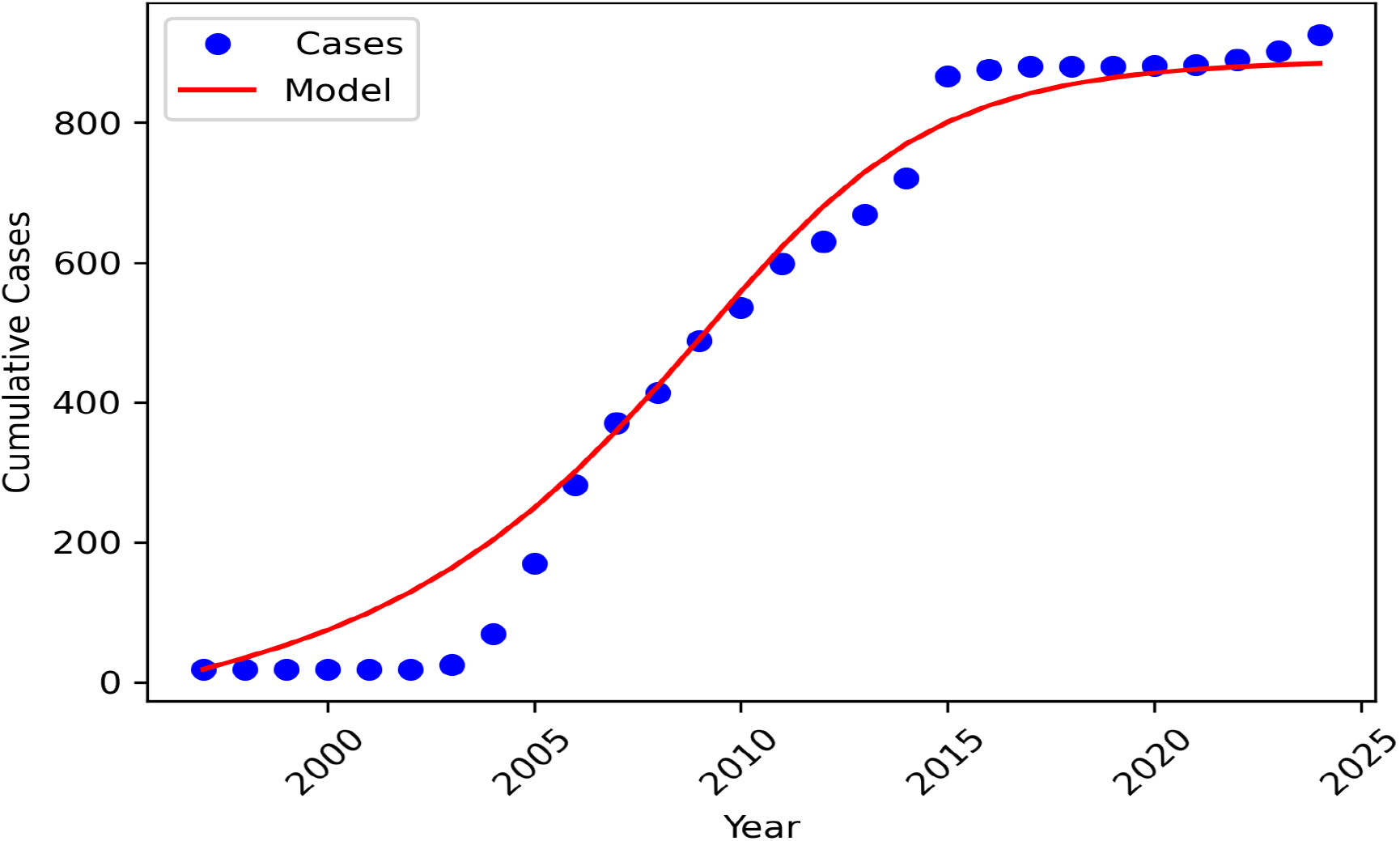
Least square model fitting on the cases data.

Figure 2.1, obtained from [26], represents the time-scaled maximum-likelihood tree of HPAI on the basis of 1, 000 sequences. The main clades are labeled red and the pink-hued bar indicates the wild-bird resurgence events. The figure shows that there are putative transmission pathways between wild birds, domestic birds, and humans. We note that the susceptible and infectious domestic and wild populations are reduced by natural death at a rate *µ*_*d*_, *µ*_*w*_. However, for domestic birds, due to mass culling of exposed and infected birds, this group is further reduced by *c*_*E*_, *c*_*I*_, which represents the culling rate of exposed and infectious domestic birds, respectively. From the defined compartments, the total human, domestic, and wild populations, denoted as *N*_*h*_, *N*_*d*_ and *N*_*w*_ are defined as

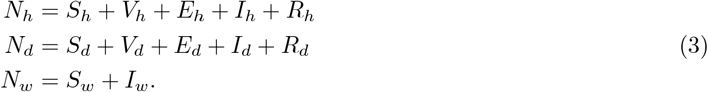

Following the outlined descriptions, the system of nonlinear ordinary differential equations (ODEs) formulated to examine the dynamics of H5N1 transmission in this study is defined by (4) with the flowchart shown in Figure 2 while the model’s state parameters and their description are given in Table 2.

**Table 2:**
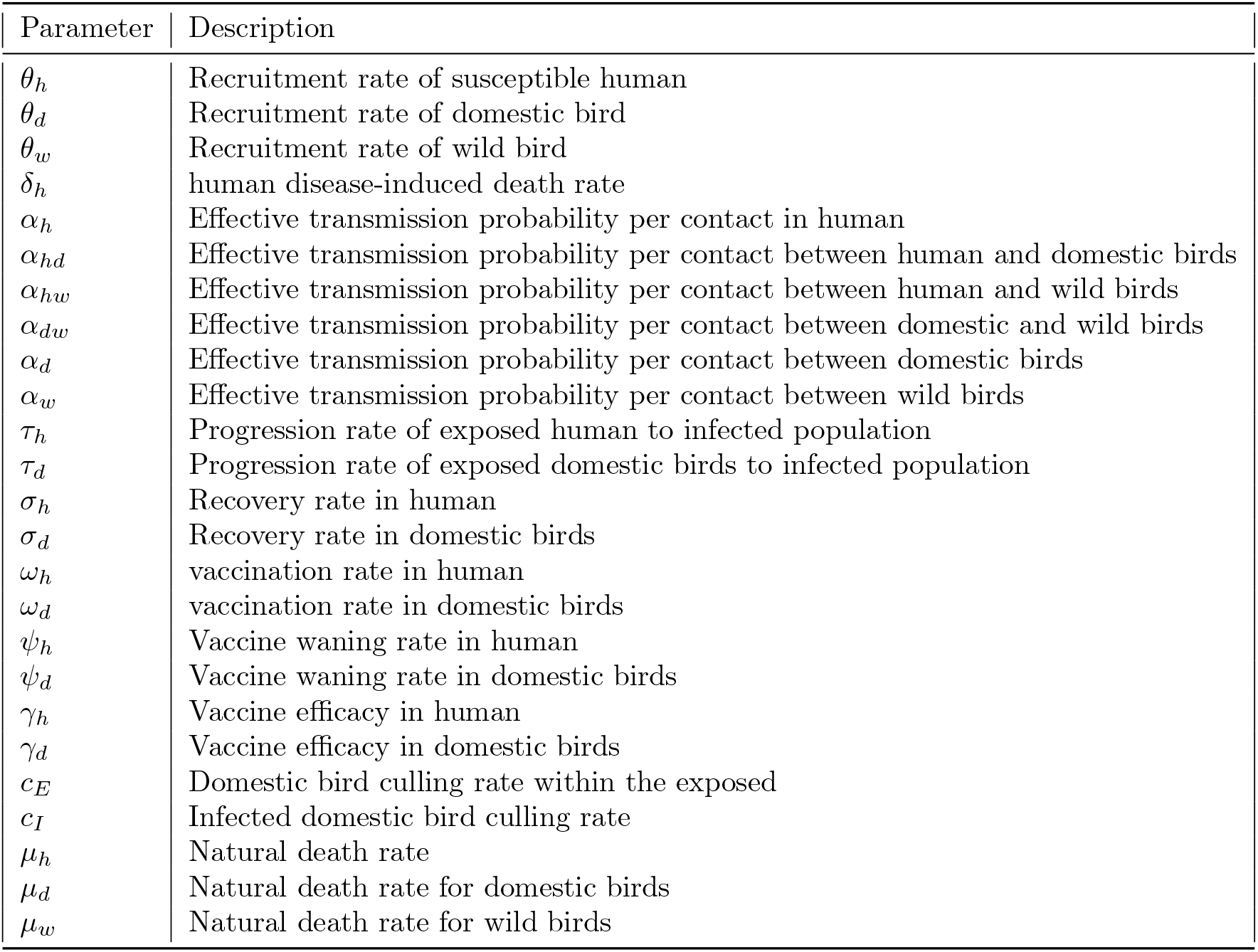
Description of model parameters.

| Parameter | Description |
| --- | --- |
| $\theta_h$ | Recruitment rate of susceptible human |
| $\theta_d$ | Recruitment rate of domestic bird |
| $\theta_w$ | Recruitment rate of wild bird |
| $\delta_h$ | human disease-induced death rate |
| $\alpha_h$ | Effective transmission probability per contact in human |
| $\alpha_{hd}$ | Effective transmission probability per contact between human and domestic birds |
| $\alpha_{hw}$ | Effective transmission probability per contact between human and wild birds |
| $\alpha_{dw}$ | Effective transmission probability per contact between domestic and wild birds |
| $\alpha_d$ | Effective transmission probability per contact between domestic birds |
| $\alpha_w$ | Effective transmission probability per contact between wild birds |
| $\tau_h$ | Progression rate of exposed human to infected population |
| $\tau_d$ | Progression rate of exposed domestic birds to infected population |
| $\sigma_h$ | Recovery rate in human |
| $\sigma_d$ | Recovery rate in domestic birds |
| $\omega_h$ | vaccination rate in human |
| $\omega_d$ | vaccination rate in domestic birds |
| $\psi_h$ | Vaccine waning rate in human |
| $\psi_d$ | Vaccine waning rate in domestic birds |
| $\gamma_h$ | Vaccine efficacy in human |
| $\gamma_d$ | Vaccine efficacy in domestic birds |
| $c_E$ | Domestic bird culling rate within the exposed |
| $c_I$ | Infected domestic bird culling rate |
| $\mu_h$ | Natural death rate |
| $\mu_d$ | Natural death rate for domestic birds |
| $\mu_w$ | Natural death rate for wild birds |

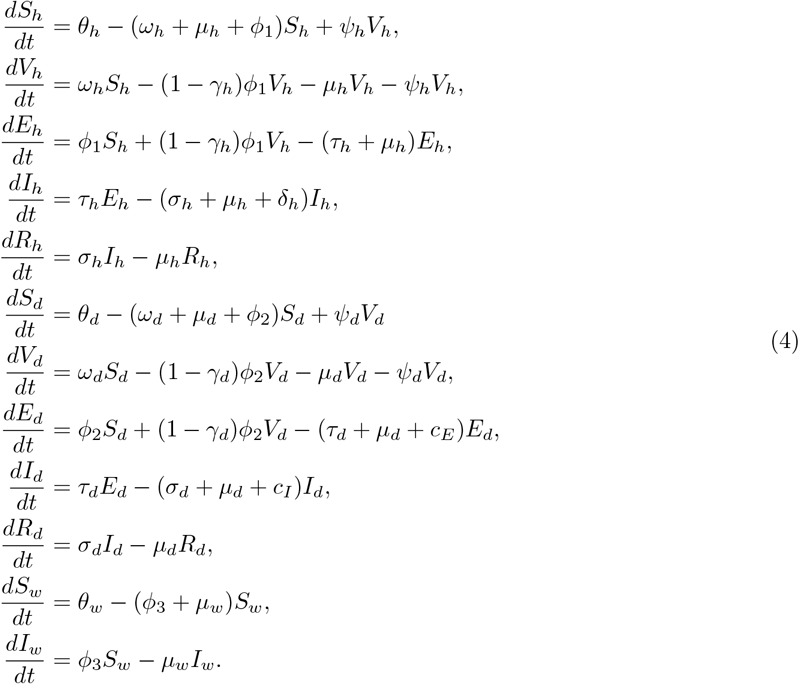

### 2.2. Model Analysis

Here, we present the analysis of the model presented in (4) by considering each group. The total population within the human, domestic, and wild bird population are denoted as *N*_*h*_, *N*_*d*_, and *N*_*w*_ respectively. For the human population, using (3), we have

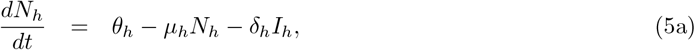

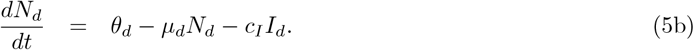

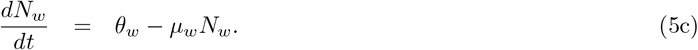

Thus, using the results from (5a)–(5c), we have the theorem that shows the positivity and boundedness of the model.

### 2.3. Positivity and Boundedness

**Theorem 1**. *Suppose* (*S*_*h*_, *V*_*h*_, *E*_*h*_, *I*_*h*_, *R*_*h*_, *S*_*d*_, *V*_*d*_, *E*_*d*_, *I*_*d*_, *R*_*d*_, *S*_*w*_, *I*_*w*_) *are the solutions of the model provided in* (4) *with the initial conditions defined in a biologically feasible region* Ω = Ω_*h*_ × Ω_*d*_ × Ω_*w*_, *where*

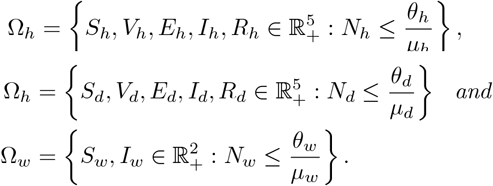

*Then, the set* Ω *is a non-negative variant*.

**Proof**. From (5a), we have

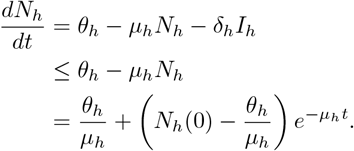

This shows that

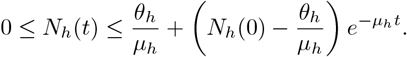

Also, from (5b), we have

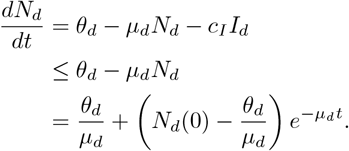

This shows that

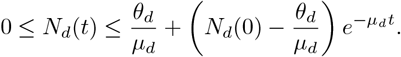

Similarly, using (5c), we obtained

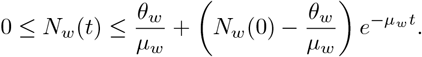

Hence, the Ω is positive invariant for all *t*. Also, as *t* −→ ∞, we have

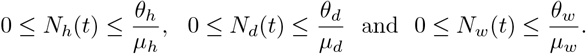

This shows that the system is positive and bounded.

### 2.4. Existence of H5N 1 disease-free equilibrium

At the *H*5*N* 1 equilibrium state of the system (4), we have

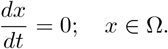

Thus the *H*5*N* 1 equilibrium free state is obtained as

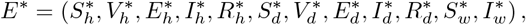

Where

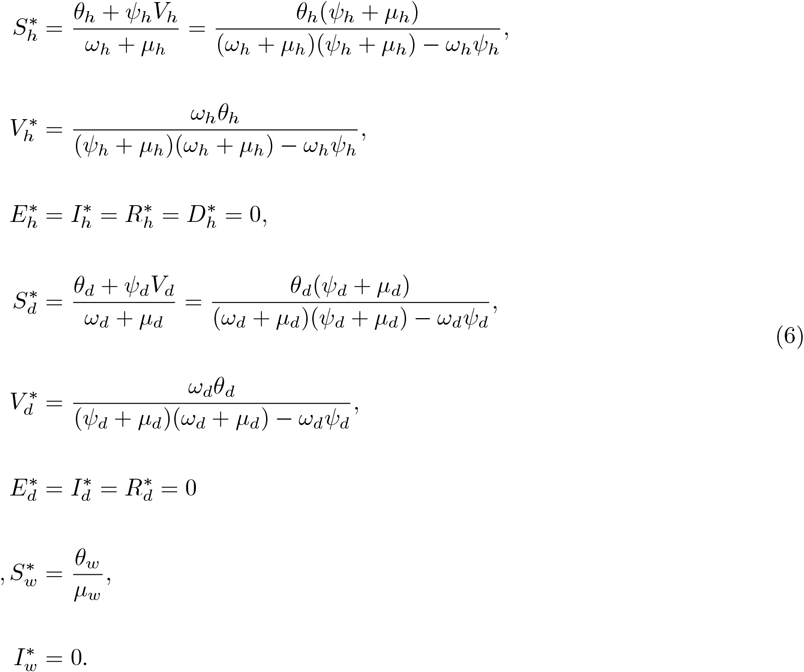

### 2.5. The basic reproduction number

The basic reproduction number often referred to as the ℛ_0_ is the fundamental concept that describes the tendency of disease spread. This quantifies the pathogen’s transmission potential within the population and represents the average number of individuals likely to become infected by a single case when introduced into a susceptible population[27]. Thus, to predict the potential impact of H5N1 and to determine both the effectiveness and necessity of the control measure, calculating the basic reproduction number is essential. As a result, we determine the basic reproduction number for the three population groups in our proposed model given in (4). We use the next-generation matrix, widely used by [19, 28–33], where the basic reproduction number is the spectral radius of the next-generation matrix. Using this approach, we assemble the two main matrices say *F* = (*F*_*h*_, *F*_*d*_, *F*_*w*_) and *V* = (*V*_*h*_, *V*_*d*_, *V*_*w*_). Here, matrix *F* represents the transmission components of the infected state of the model and *V* describes the transition between and out of the infected states. In our model, the transmission components are the exposed (*E*_*h*_, *E*_*d*_) and infected (*I*_*h*_, *I*_*d*_, *I*_*w*_). We denote the basic reproduction number within the human population as 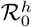 while domestic and wild birds are represented by 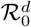 and 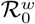, respectively. For human, we have

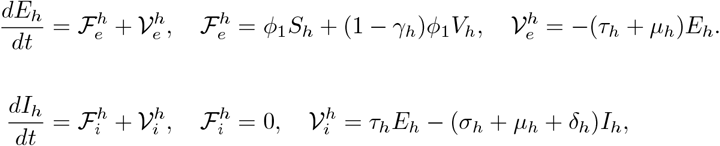

Hence, the matrix *F*_*h*_ and *V*_*h*_ are given as

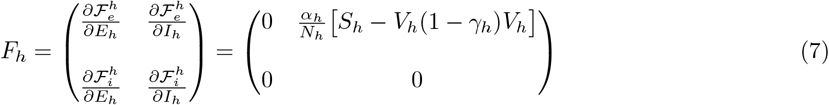

and

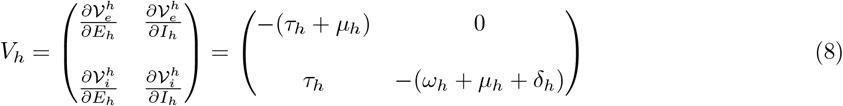

Hence, the next-generation matrix of the human population (say *M*_*h*_) is given by 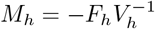. That is,

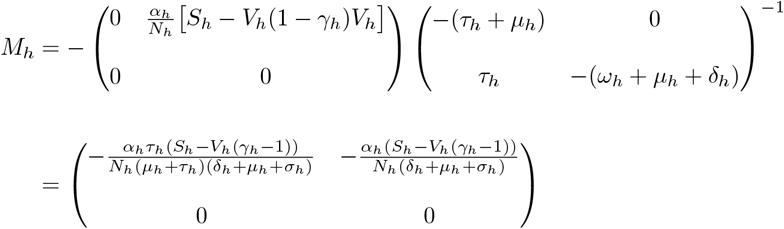

Thus, 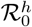 which is the spectral radius of the next-generation matrix is given as 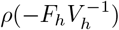. Therefore,

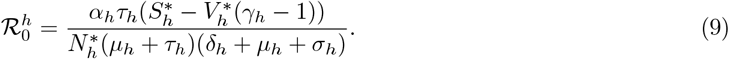

Similarly, the basic reproduction number for both domestic birds are obtained as:

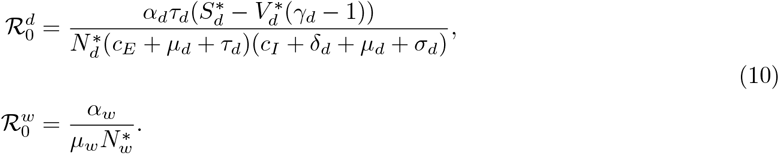

Thus,

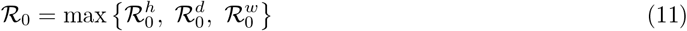

We could also obtain the the model R_0_ directly as a combined group where we will have

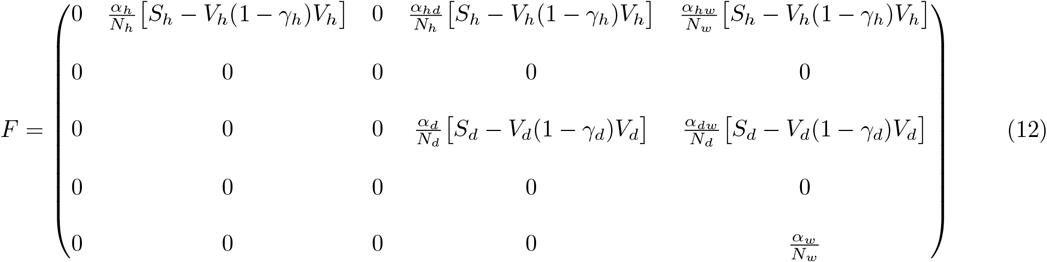

and

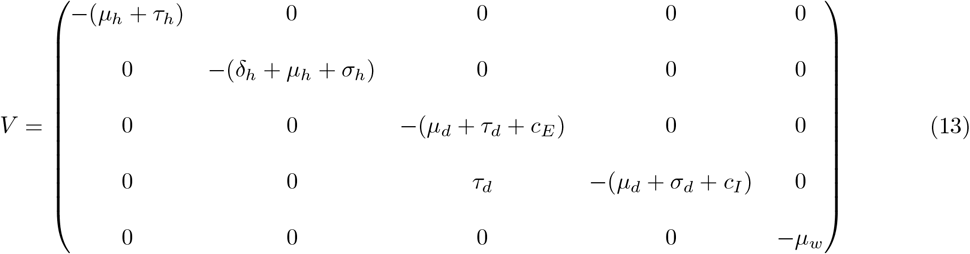

Hence,

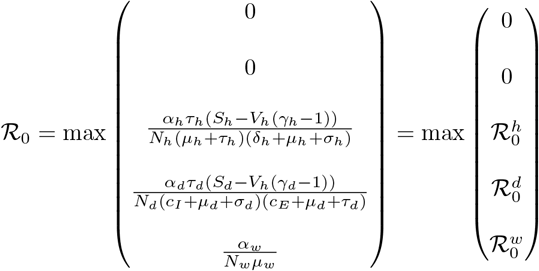

If ℛ_0_ *<* 1, then this implies that the disease dies out within the population, while ℛ_0_ *>* 1 implies that the disease continues within the population suggesting a possible endemic state. However, to determine which transmission parameters have a significant influence on the ℛ_0_, we calculate the ℛ_0_ with varying parameters. Thus, this will lead us to the sensitivity analysis which helps to determine the relationship between ℛ_0_ and the parameters.

### 2.6. Model fitting and parameter estimations

In this section, we estimate the model parameter values provided in the model (4). This estimate enables us to determine the key driver of disease spread and identify the base values. In this work, we adopted the least square method for our parameter estimation. It is understood that the least square method is a powerful tool for parameter estimation as it helps to find the best curve representing the data and minimize the error between the model and the data points [32, 34]. Our data fitting was conducted using publicly available H5N1 human case data obtained from the World Health Organization [35] and also documented in [36]. We note that some parameters are assumed while some are estimated from the literature. For the parameter within the domestic bird component, the existing parameters in the literature are used as guidance, which are then calibrated into our model for validation.

For the least square fit, the objective function to be minimized is the root mean square error defined by:

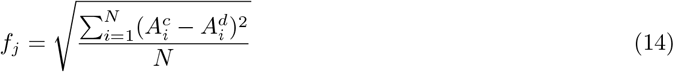

where 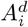 represents the recorded cumulative cases of infected individuals within the period of consideration of length *N* while 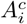 is the corresponding model output. In this estimation, we used the incidence data for the global case data from 1997 to 2025. The incidence data and the model fitting curve with the estimated parameter values are shown in Figure 2.6.

Hence, the obtained parameter values are given in Table 3

**Table 3:** Description of model parameters and the estimated value.

| Parameter | Description | Parameter range | Baseline Value | Source |
| --- | --- | --- | --- | --- |
| $\theta_h$ | Recruitment rate of susceptible human | 0.1 – 0.5 | 0.479 | Fitted |
| $\theta_d$ | Recruitment rate of domestic bird | 0.01 – 0.1 | 0.1 | Estimated [37, 38] |
| $\theta_w$ | Recruitment rate of wild bird | 0.1 – 0.6 | 0.524 | Estimated [39, 40] |
| $\delta_h$ | human disease-induced death rate | 0.01 – 0.5 | 0.0474 | Fitted |
| $\alpha_h$ | Effective transmission probability per contact in human | 0.0 – 0.3 | 0.30 | Assumed [41] |
| $\alpha_{hd}$ | Effective transmission probability per contact between human and domestic birds | 0.4 – 0.6 | 0.48 | Fitted |
| $\alpha_{hw}$ | Effective transmission probability per contact between human and wild birds | 0.5 – 0.7 | 0.59 | Fitted |
| $\alpha_{dw}$ | Effective transmission probability per contact between domestic and wild birds | 0.5 – 0.6 | 0.51 | Fitted |
| $\alpha_d$ | Effective transmission probability per contact between domestic birds | 0.4 – 0.5 | 0.441 | Fitted |
| $\alpha_w$ | Effective transmission probability per contact between wild birds | 0.3 – 0.7 | 0.432 | Estimated [42–45] |
| $\tau_h$ | Progression rate of exposed human to infected population | 0.7 – 0.9 | 0.896 | Fitted |
| $\tau_d$ | Progression rate of exposed domestic birds to infected population | 0.1 – 0.5 | 0.456 | Estimated [46] |
| $\sigma_h$ | Recovery rate in human | 0.001 – 0.005 | 0.0039 | Fitted |
| $\sigma_d$ | Recovery rate in domestic birds | 0 – 0.1 | 0.0813 | Estimated [47] |
| $\omega_h$ | vaccination rate in human | 0.5 – 0.9 | 0.527 | Estimated [48] |
| $\omega_d$ | vaccination rate in domestic birds | 0.5 – 0.9 | 0.50 | Estimated [48] |
| $\psi_h$ | Vaccine waning in human | 0.4 – 0.9 | 0.415 | Assumed [49, 50] |
| $\psi_d$ | Vaccine waning rate in domestic birds | 0.3 – 0.6 | 0.422 | Assumed [51, 52] |
| $\gamma_h$ | Vaccine efficacy in human | 0.1 – 0.6 | 0.54 | Assumed [53] |
| $\gamma_d$ | Vaccine efficacy in domestic birds | 0.5 – 0.9 | 0.60 | Assumed [24] |
| $c_E$ | Domestic bird culling rate within the exposed | 0.01 – 0.5 | 0.0416 | Assumed [54] |
| $c_I$ | Infected domestic bird culling rate | 0.5 – 1.0 | 0.50 | Assumed [54] |
| $\mu_h$ | Natural death rate | 0.1 – 0.5 | 0.298 | Fitted |
| $\mu_d$ | Natural death rate for domestic birds | 0.01 – 0.1 | 0.0936 | Estimated [55–57] |
| $\mu_w$ | Natural death rate for wild birds | 0.1 – 0.3 | 0.117 | Estimated [58–60] |

### 2.7. Sensitivity analysis

The sensitivity analysis is a crucial tool used to evaluate how the uncertainty in the model output can be attributed to its inputs by varying the parameters and observing the resulting changes. This tool helps to identify key variables with the most significant impact on the system behavior, which thereby enhances the understanding of their relationships. With respect to the R_0_, we observed that reproduction number given in (11) is a function of the following parameters: *θ, α, ω, ψ, τ*_*h*_, *σ, µ, δ, c*_*E*_, *c*_*I*_. So, we determine the strength of their relationship using the partial rank correlation coefficient, and the results are displayed in Figure 4.

**Figure 4:**
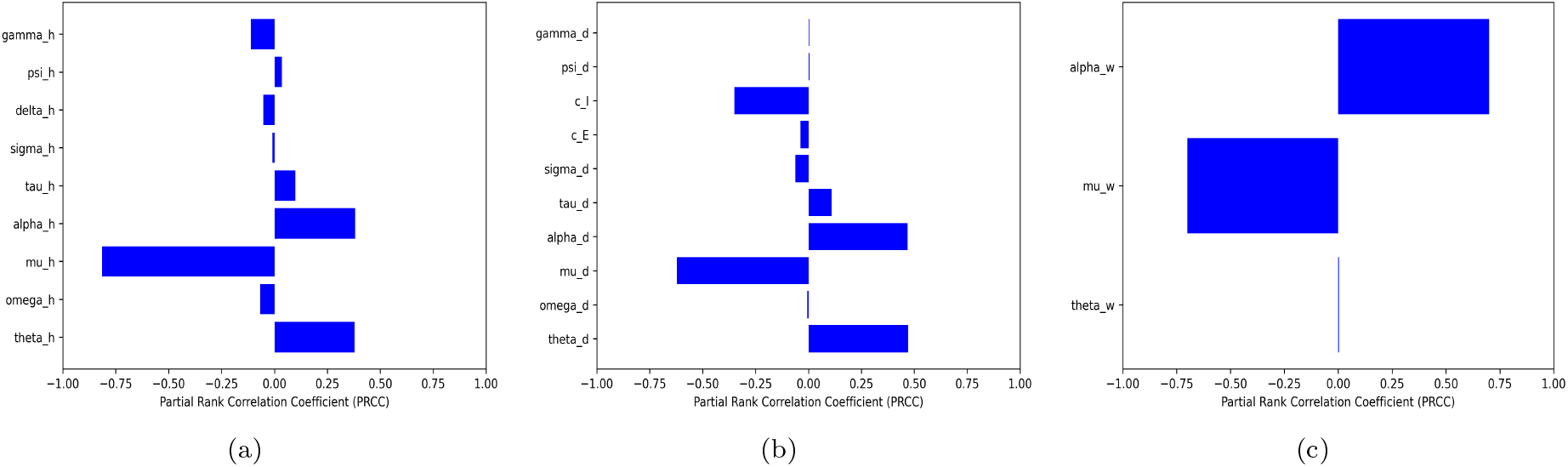
Sensitivity analysis (a) Human (b) Domestic (c) Wild.

From Figure 4, we can observe that there are positive and negative relationships between the parameters and ℛ_0_. For instance, the figures suggest that ℛ_0_ has a positive relationship with an increase in transmission probability and a negative relationship when there is an increase in vaccine coverage and efficacy. However, the disease progression rate, transmission probability, and recovery rate appear to have a more significant impact on the ℛ_0_ while vaccination rate and efficacy show a slight influence. It is important to note that this does not suggest that the vaccination rate and the efficacy are not essential for reducing ℛ_0_. On the contrary, it merely suggests that each parameter alone might not provide a significant impact mitigating the spread. As such, we shall further present the pair variation of these parameters with the ℛ_0_ to determine the relationship.

As seen in Figure 5 representing the variation of the basic reproduction number corresponding to varying pair parameters. The figure shows that the increment of the vaccine efficacy and the coverage significantly reduces ℛ_0_. So, this confirms that high vaccine efficacy with above 50% vaccination rate in humans would be necessary to maintain ℛ_0_ *<* 1.

**Figure 5:**
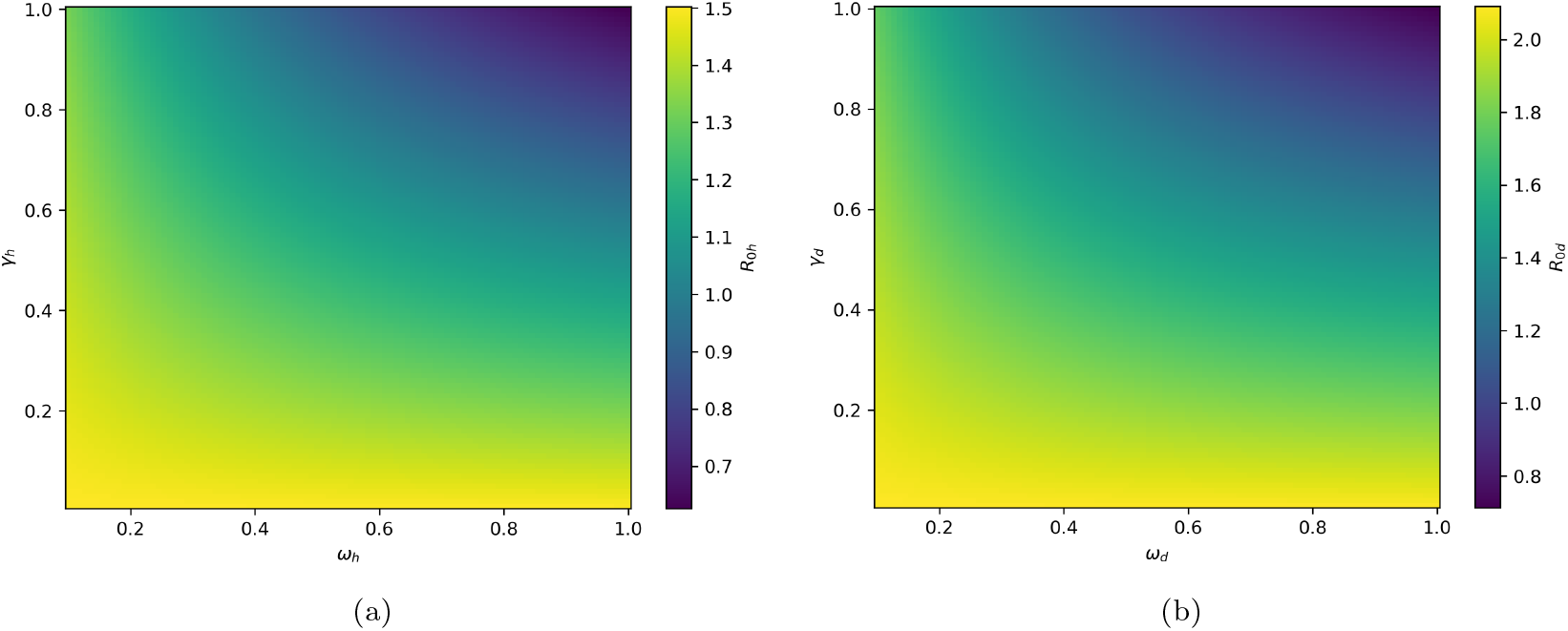
Pair vaccination rate and vaccine efficacy heat map (a) Human (b) Domestic.

## 3. Results

### 3.1. Numerical Simulation

We performed a numerical simulation of the model presented in (4) using the baseline parameter provided in Table 3 to assess the impact of cross-species transmission per contact rate and vaccine efficacy on the spread of the H5N1 in humans. The results are presented in Figure 6.

**Figure 6:**
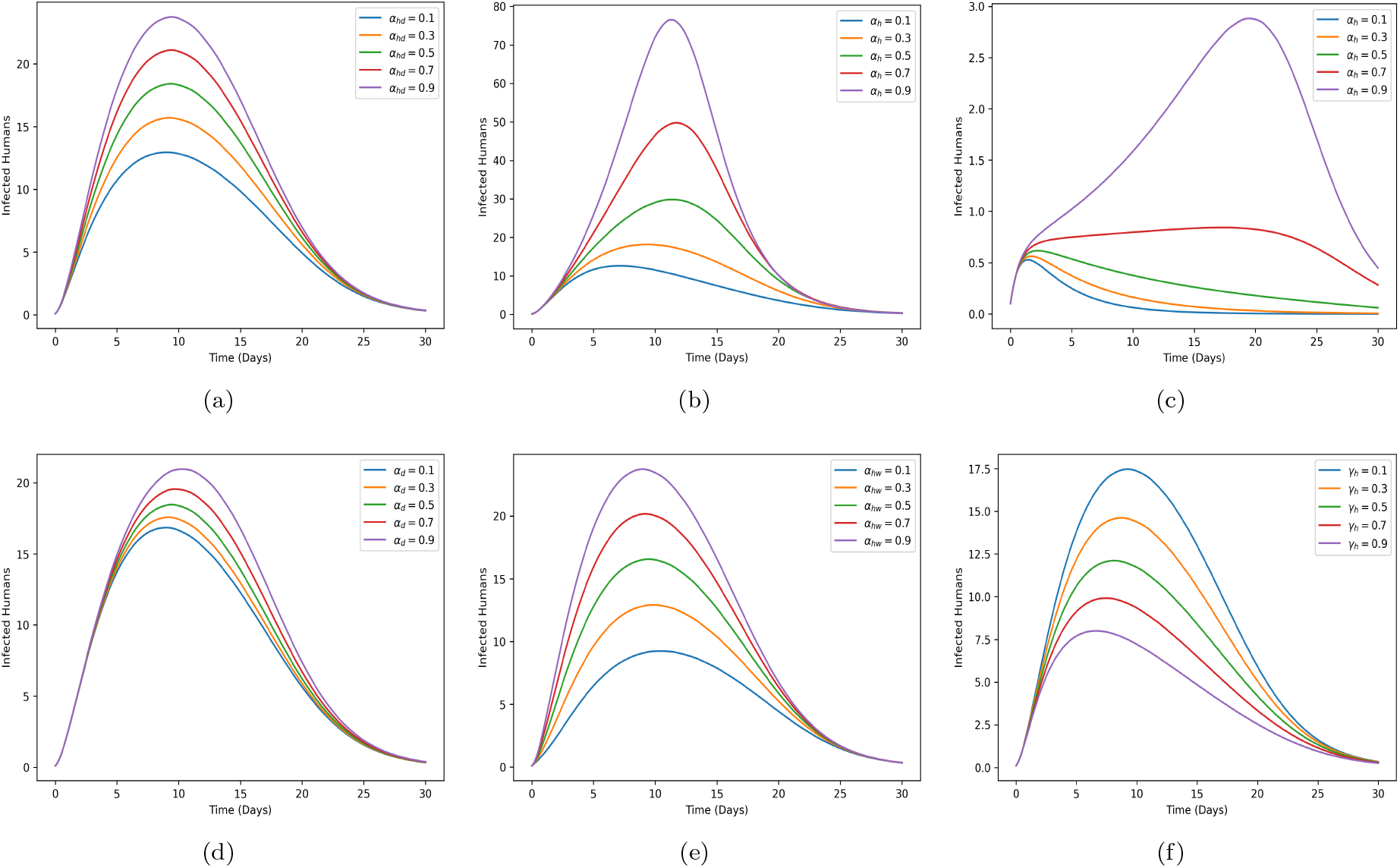
Effective per contact rate (a) Domestic-Human (b) Within Human (c) Within Human with no other transmission route (d) Domestic-Domestic (e) Wild-Human (f) Effect of varying vaccine efficacy.

The results shown in Figure 6, highlight that effective transmission per contact between each species has a significant impact on disease transmission and, as this parameter decreases, there is a reduction in the number of new infections over time. Moreover, Figure 6(c) shows that the better way to have the least spread of the virus in humans might not necessarily be to focus on humans, but rather to tackle the natural host and the main transmission pathway which would be to focus on domestic and wild birds. This suggests that if the virus could be controlled in animals and we would limit the interaction between humans and infected domestic and wild birds, then the virus can be contained to a bare minimum in humans within 30 days.

### 3.2. Optimal control strategy

In this section, we perform the optimal control strategy and implement the four time-dependent control variables to explore their effectiveness in controlling the spread. Our proposed control variables are represented by *u*_1_(*t*), *u*_2_(*t*), *u*_3_(*t*) and *u*_4_(*t*). These variables are defined below:

- *u*_1_(*t*) represents the environmental sanitation. This is considered as the measure put in place to prevent the susceptible from being exposed or infected. This measure mainly targets the susceptible population that is more at risk for infection and could further include personal hygiene and the use of personal protective equipment.
- *u*_2_(*t*) represents early vaccination strategy. It is presumed that there are vaccines available to be administered, and mainly focuses on the vaccination compartment within the model to ensure that there is a level of defense against the virus.
- *u*_3_(*t*) represents the early treatment strategy for infected individuals that incorporates early detection, testing, and quarantine. The goal of this strategy is to determine whether early treatment could help in recovery and limit long-term spread in humans.
- *u*_4_(*t*) represents the culling strategy within the domestic bird population. This strategy is already adopted across many regions. So, the objective is to determine the effectiveness of this strategy within domestic birds.

With the implementation of the above strategy, the result of the optimal control model is provided in the Figures 7 - 9. The formulation of the optimal model well documented in the Appendix

**Figure 7:**
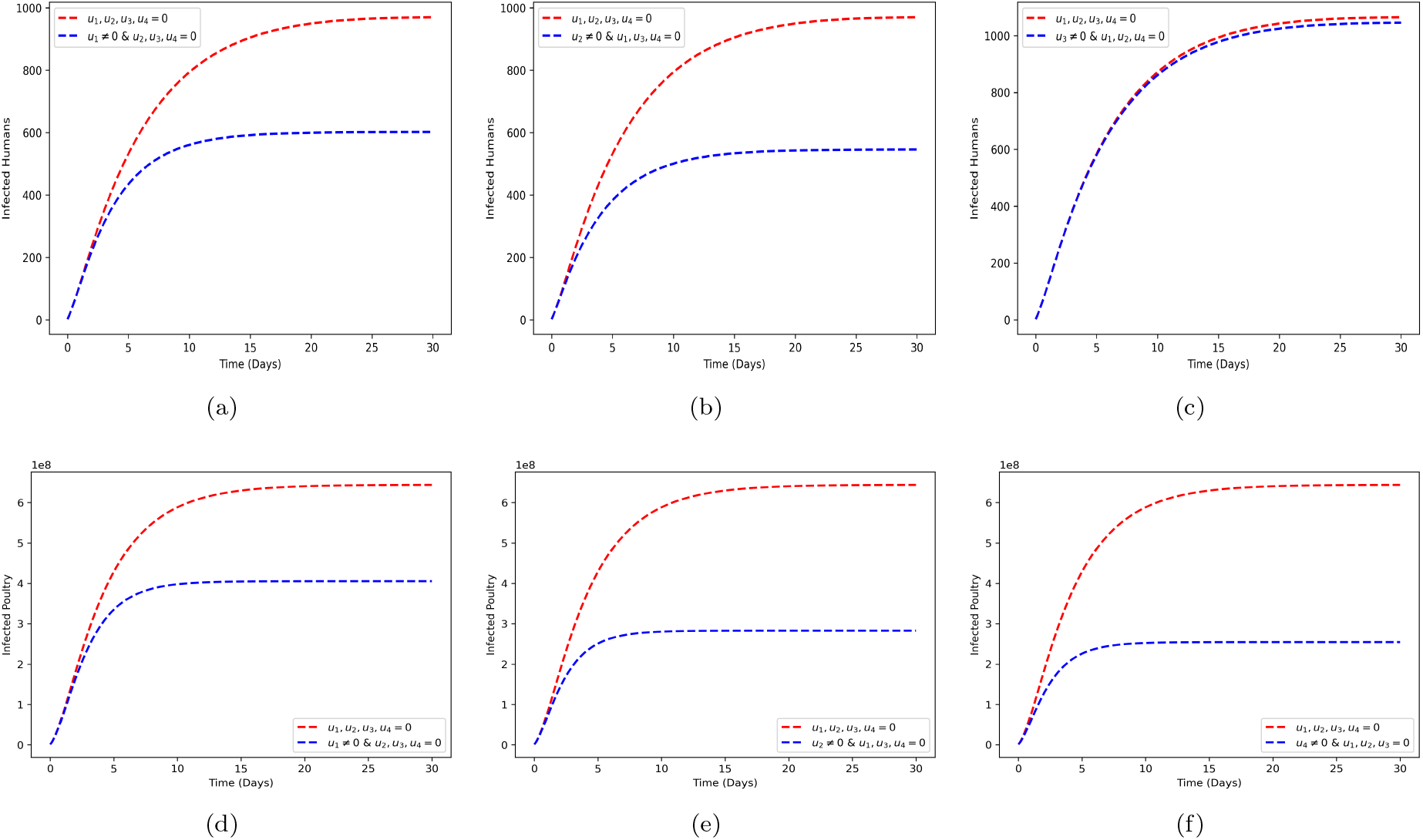
Implementation of a single control measure in both human and poultry.

Figure 7 shows the result of implementing a single control measure. Figures 7(a) and 7(d) represent the implementation of environmental sanitation in humans and poultry, respectively, while 7(b) and 7(e) represent the implementation of vaccination in humans and poultry. Finally, 7(c) and 7(f) represent the implementation of early treatment in humans and culling in poultry, respectively. By comparing these measures, the result shows that vaccination as a single measure works best in humans while culling provides the highest averted cases in poultry. Next, we show the implementation of double control measures which are shown in Figure 8.

**Figure 8:**
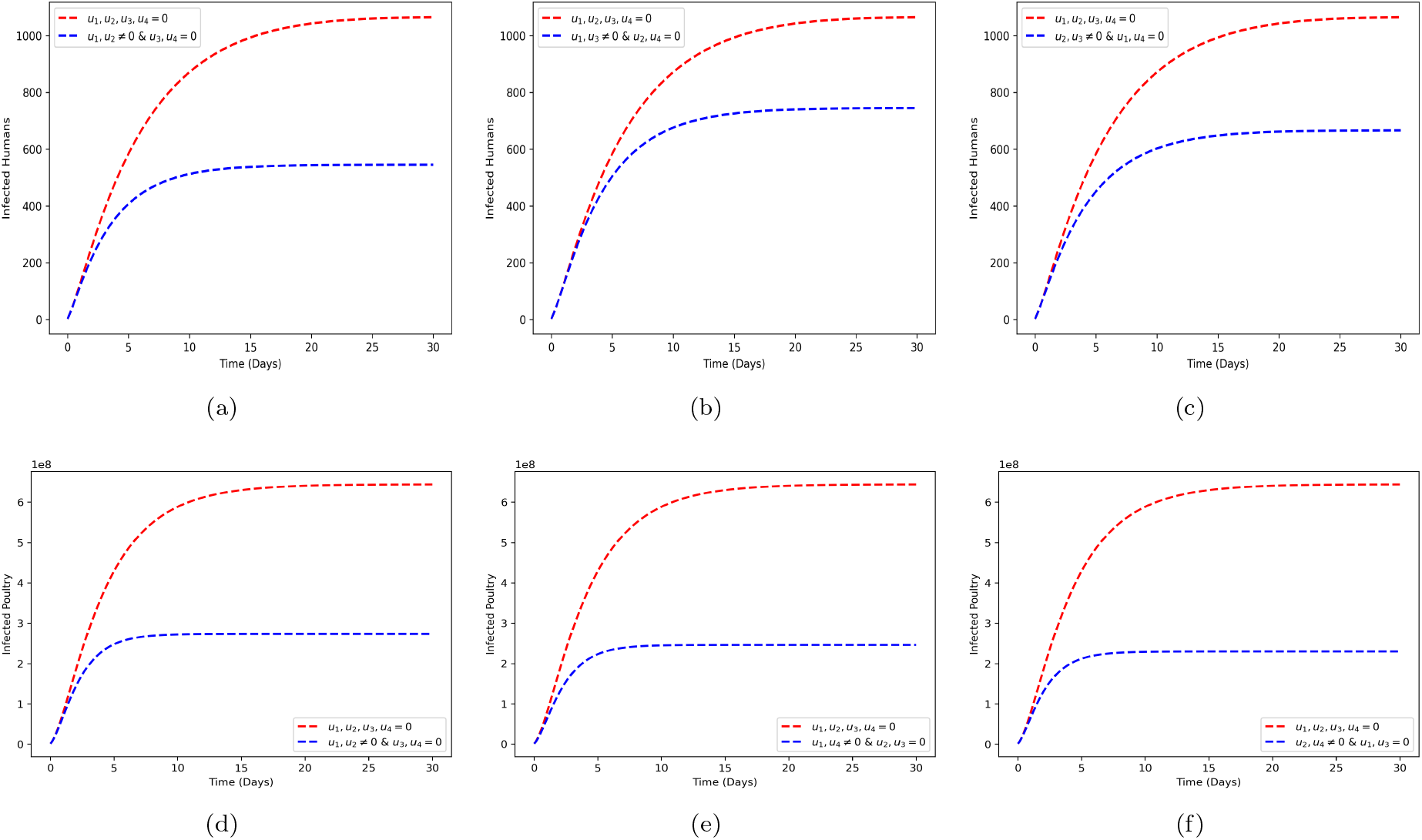
Implementation of a double control measure in both human and poultry.

The implementation of a double control measure further reduces infection in both humans (8(a), 8(b), 8(c)) and poultry (8(d), 8(e), 8(f)). In particular, the combination of environmental sanitation and vaccination do best in humans, while the combination of vaccination and culling of infected poultry show the least new infection in poultry. Finally, we show the spread of the disease under the implementation of all combined control measures.

As shown in Figure 9, implementing the combined measures further reduces the number of infections in humans and poultry. Therefore, it is suggested that the combination of the proposed control measure needs to be implemented to have the best control over the outbreak. In general, results show the impact of several control measures on both human and poultry populations. The figures outline the outcome for implementing single, double, and combined strategies, respectively. As observed in the figures, the implementation of the three strategies (*u*_1_, *u*_2_, *u*_3_) provides more protection against spread compared to the implementation of the double control strategies (*u*_1_, *u*_2_). Similarly, we observe that there are fewer cases obtained in poultry when all strategies (*u*_1_, *u*_2_, *u*_4_) are implemented.

**Figure 9:**
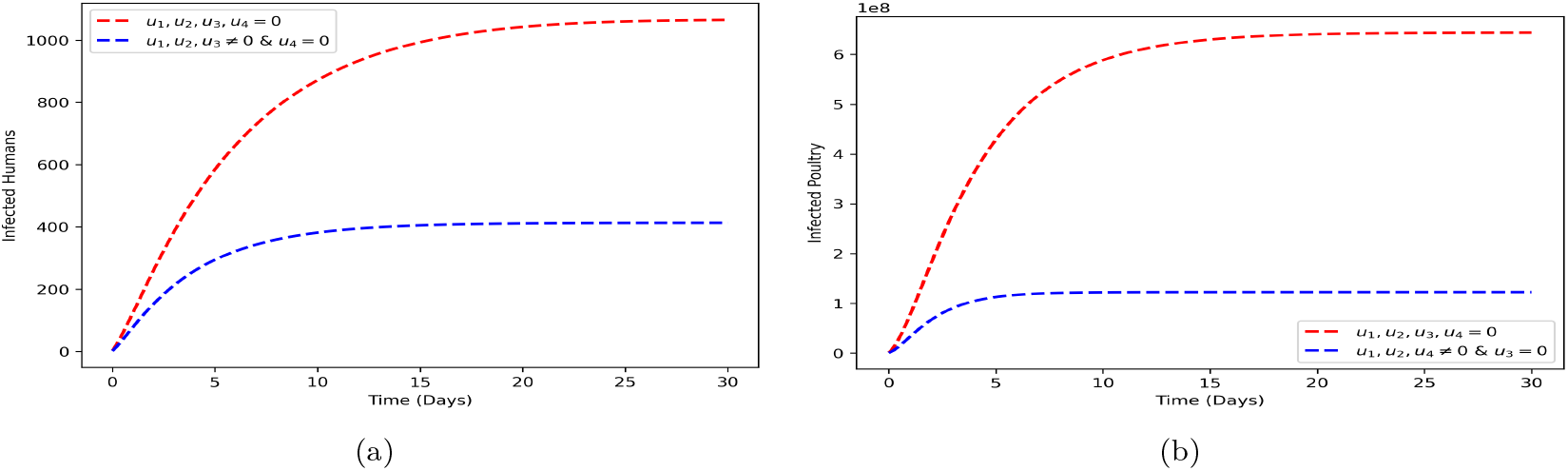
Implementation of all control measure in both human and poultry.

## 4. Discussion and Conclusion

In this work, we adopted a phylogenetic analysis and leveraged its findings to develop a mathematical model that examines the transmission dynamics of H5N1 between humans, domestic poultry, and wild birds. Using this model, we derived the basic reproduction number and found that high vaccine efficacy combined with at least 50% vaccination coverage is required to have the ℛ_0_ *<* 1 which is crucial for controlling the virus in human populations. In addition, the sensitivity analysis highlighted the importance of inter-population transmission, particularly from wild to domestic birds, in sustaining outbreaks. The findings suggest, that human infections are primarily driven by contact with infected domestic poultry rather than direct spillover from wild birds. This emphasizes the need for better disease management within poultry farms to reduce spillover risks to humans. We also found that targeted interventions in poultry populations could significantly reduce human infections. In addition, it is believed that by implementing strict biosecurity measures, such as improved sanitation and farm hygiene protocols, we could further mitigate disease spread. To assess the effectiveness of the intervention strategies, we performed an optimal control analysis, which evaluates four key measures: environmental sanitation, vaccination, early treatment, and targeted culling of infected poultry. Our results indicate that a combination of all the strategies yields the greatest reduction in disease transmission for both humans and poultry. However, from a effectiveness perspective, the optimal approach varies:

1. For human populations, a combination of environmental sanitation and vaccination is sufficient to mitigate spread, as the treatment measure has less of an effect on the number of cases averted.
2. For poultry, a strategy involving environmental sanitation with vaccination and targeted culling is proven to provide the best protection.

Furthermore, the results show that environmental sanitation plays an important role in the control of the outbreak, since this alone reduces the prevalence of infection in poultry and would minimize the tendency of spillover in humans. Therefore, it is important to pay more attention to the environment. In general, these findings reinforce the importance of environmental sanitation in reducing indirect transmission through contaminated surfaces and emphasize the need for hygiene practices in human, poultry and live bird markets. Despite the strengths of our model, we acknowledge certain limitations. Our model assumed instantaneous transitions between disease states, which may not always hold, especially in cases of asymptomatic infections where there could be a delay before infection and testing. Therefore, this work might be improved by incorporating spatial heterogeneity to better reflect a more accurate transmission pattern and inclusion of time delays in disease progression. Finally, while we integrated phylogenetic understanding to inform spillover events, further validation using surveillance data, seasonal variations in wild bird migration, stochastic effects, and adaptive human behavioral responses would enhance the robustness of future models. To address some of these limitations, a more detailed inter-species transmission model is being developed by [61], which incorporates both wild and domestic mammals and leverages genomic sequence data through Bayesian phylogenetic inference using the PhyDyn package for parameter estimation. This approach aims to improve inference on cross-species transmission dynamics and provide a more precise quantification of ecological processes influencing disease spread. In conclusion, this work underscores the need for a multifaceted approach to managing H5N1 outbreaks. Through the integration of mathematical modeling, phylogenetic analysis, and control evaluations, we provided a framework to support the design of optimal intervention strategies that could balance disease control and economic feasibility.

## Data Availability

All data used are publicly available online

## Data and Code Availability

All data used in this work are publicly available

### Competing interest

Authors declare no conflict of interest

### Author Contributions

O.B., M.B., and J.B. contributed to the conceptualization and design of the study, including the formulation of the research questions and modeling framework. O.B. led the development of the methodology and was responsible for writing the original draft of the manuscript. O.B., M.B., C.N.N., S.S., M.M and J.B contributed to critical review and editing of the manuscript to ensure clarity, accuracy, and scientific integrity. O.B., S.S., and M.M. performed data analysis, model implementation, and visualization of the results. All authors read and approved the final version of the manuscript.

### Funding

This work has been funded in part from the National Institute of Allergy and Infectious Diseases, a component of the NIH, Department of Health and Human Services, under contract no. 75N93021C00018 (NIAID Centers of Excellence for Influenza Research and Response, CEIRR) and Centers for Disease Control and Prevention, Department of Health and Human Services, under contract NU50CK000626.

## Appendix

The optimal control model for the proposed intervention strategies is given as:

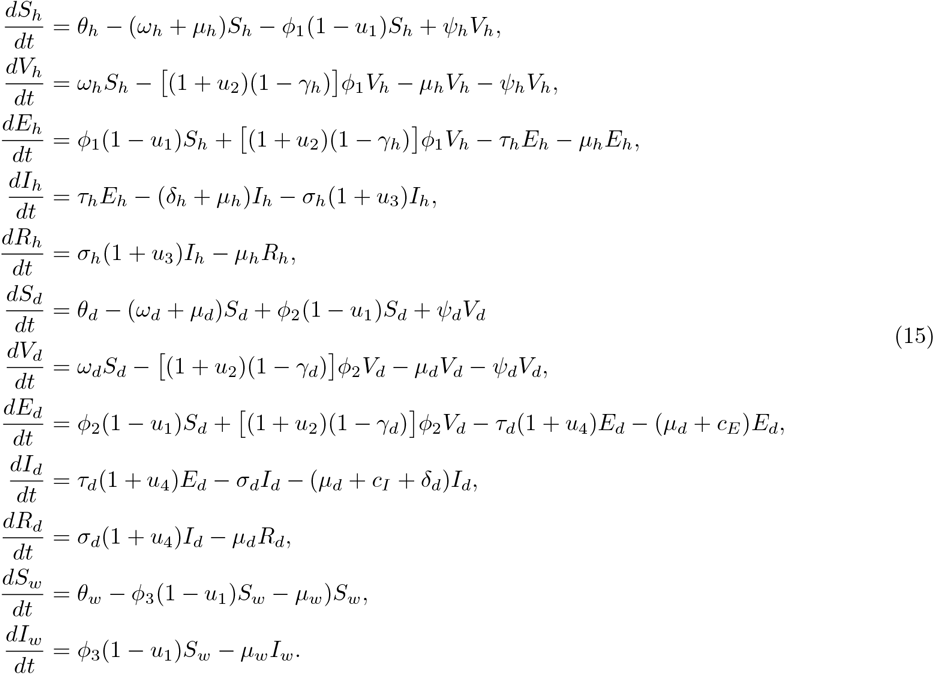

The main objective for including the control variables is to determine the optimal solution required to minimize the number of exposed and infected cases. Hence, the objective function for this optimal control problem is given by

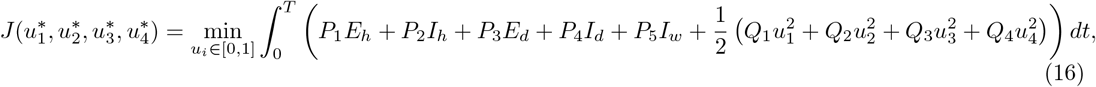

where the *P*_*i,i*=1,2,3,4,5_ are the positive weights required to balance the objective function. The linear sum *P*_1_*E*_*h*_ + *P*_2_*I*_*h*_ + *P*_3_*E*_*d*_ + *P*_4_*I*_*d*_ + *P*_5_*I*_*w*_ is the infection cost, while the quadratic sum 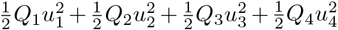 is the control cost. In particular, 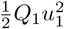 is the cost for implementing control *u*_1_ over a finite interval [0, *T*]. A similar analogy applies to *u*_2_, *u*_3_ and *u*_4_, respectively. We define the optimal control strategy 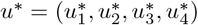 such that

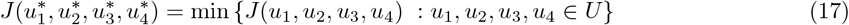

where 𝒰 is a nonempty control set defined as

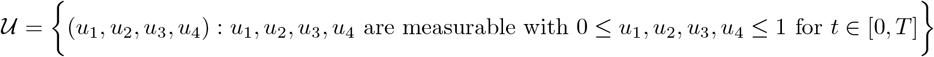

We need to adjust the necessary condition that the optimal control strategy 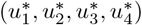 must satisfy the Pontryagin’s maximum principle [62], which changes into the optimal control problem (17) subject to the control model (15) that minimizes point-wise a Hamiltonian *H* with respect to the control measures. This Hamiltonian is given as

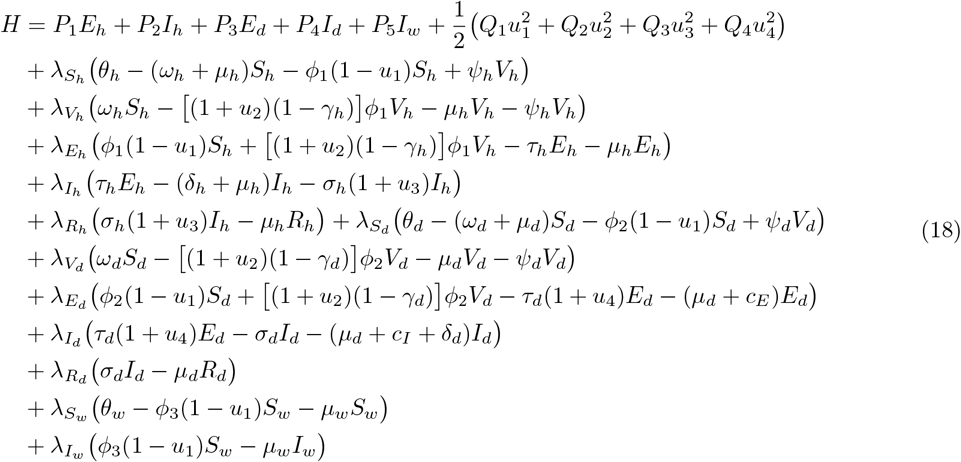

where *λ*_*i*_, *i* = *S*_*h*_, *V*_*h*_, *E*_*h*_, *I*_*h*_, *R*_*h*_, *S*_*d*_, *V*_*d*_, *E*_*d*_, *I*_*d*_, *R*_*d*_, *S*_*w*_, *I*_*w*_ represent the adjoint variables corresponding to the state variables of the model (15). Again, using Pontryagin’s maximum principle, we have the following theorem.

**Theorem 2**. *Given that* 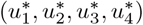 *minimizes the objective function* (16) *subject to the control model* (15), *the adjoint variables S*_*h*_, *V*_*h*_, *E*_*h*_, *I*_*h*_, *R*_*h*_, *S*_*d*_, *V*_*d*_, *E*_*d*_, *I*_*d*_, *R*_*d*_, *S*_*w*_, *I*_*w*_ *satisfy the following system*.

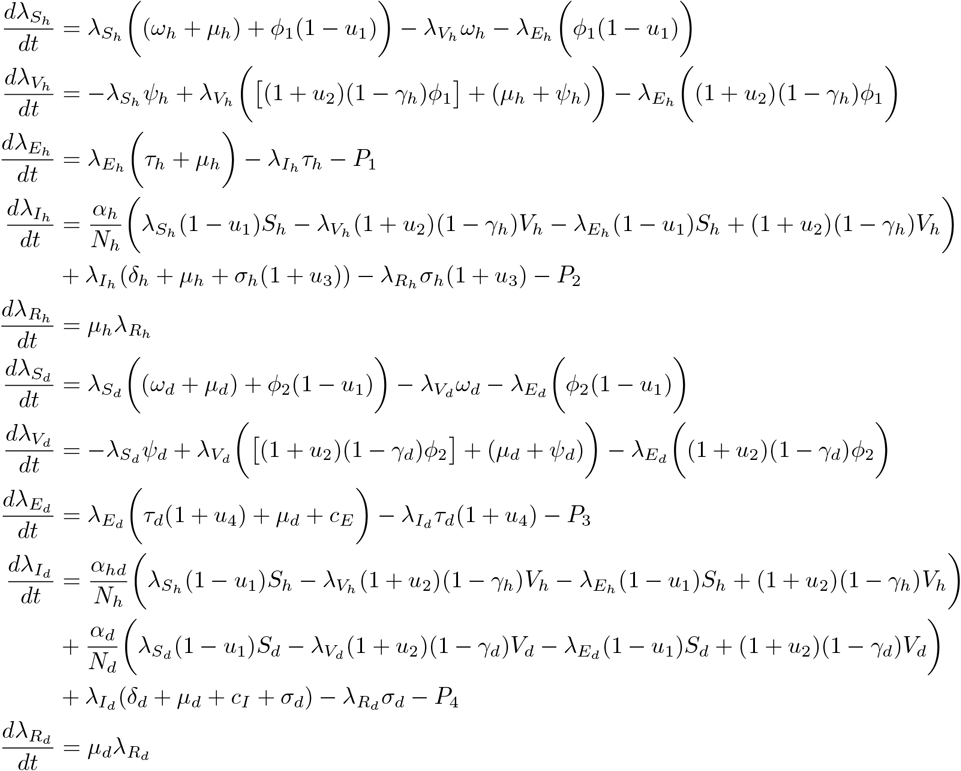

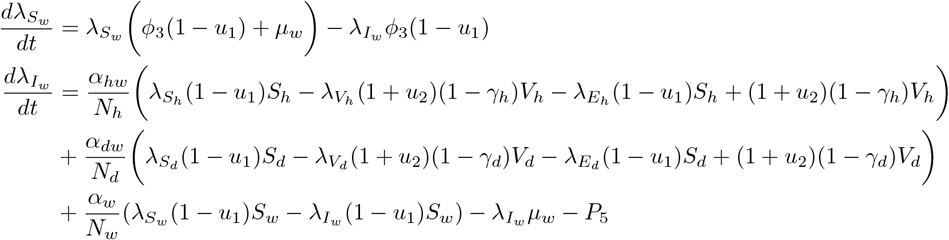

*with the terminal conditions*

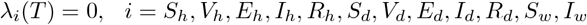

*Furthermore, the optimal control pair* 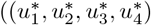 *is given as*

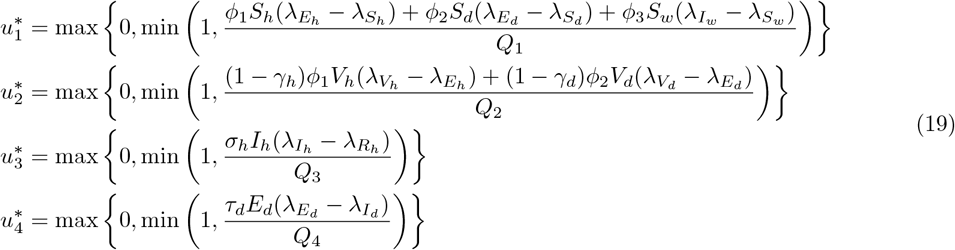

**Proof**. By differentiating the Hamiltonian *H* with respect to the state variables, we have

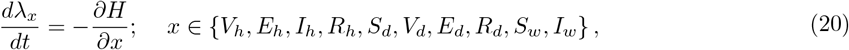

with *λ*_*i*_ = 0, for *S*_*h*_, *V*_*h*_, *E*_*h*_, *I*_*h*_, *R*_*h*_, *S*_*d*_, *V*_*d*_, *E*_*d*_, *I*_*d*_, *R*_*d*_, *S*_*w*_, *I*_*w*_. Similarly, the optimal control 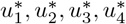 are obtained by the following optimality conditions given as

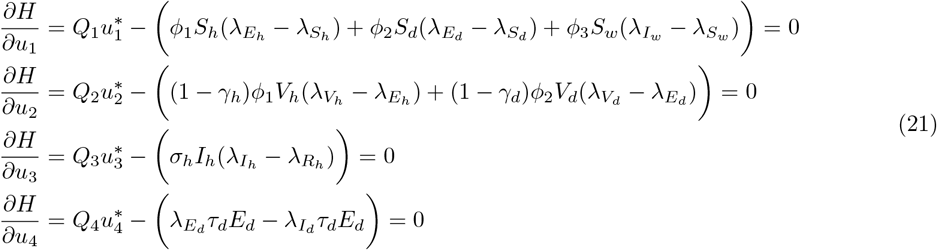

at 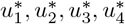 on the set 𝒰, we have

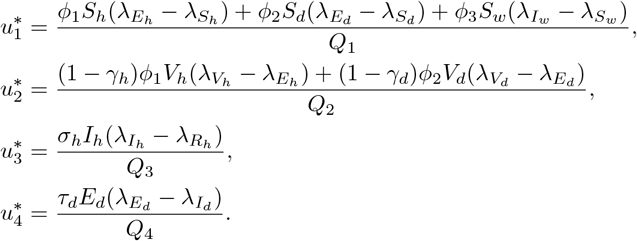

## Notes

### Competing Interest Statement

The authors have declared no competing interest.

